# The Impacts of Oral Anticoagulants on Clinical Outcomes in Patients with Atrial Fibrillation Across Five Stages of Renal Function

**DOI:** 10.1101/2024.04.15.24305865

**Authors:** Jo-Nan Liao, Yi-Hsin Chan, Hsin-Fu Lee, Yung-Hsin Yeh, Shang-Hung Chang, Shih-Ann Chen, Tze-Fan Chao

**Affiliations:** Division of Cardiology, Department of Medicine, Taipei Veterans General Hospital, Taipei, Taiwan; Institute of Clinical Medicine, and Cardiovascular Research Center, National Yang Ming Chiao Tung University, Taipei, Taiwan; The Cardiovascular Department, Chang Gung Memorial Hospital, Linkou, Taoyuan 33305, Taiwan; College of Medicine, Chang Gung University, Taoyuan 33302, Taiwan; Microscopy Core Laboratory, Chang Gung Memorial Hospital, Linkou, Taoyuan 33305, Taiwan; Graduate Institute of Clinical Medical Sciences, College of Medicine, Chang Gung University, Taoyuan, Taiwan; Center for Big Data Analytics and Statistics, Chang Gung Memorial Hospital, Taoyuan, Taiwan; Cardiovascular Center, Taichung Veterans General Hospital, Taichung, Taiwan

**Keywords:** atrial fibrillation, chronic kidney disease, Cockcroft-Gault (CG) equation, oral anticoagulant, non-vitamin K antagonist oral anticoagulant

## Abstract

**Background:** To analyze the impact of using different renal function equations and stroke prevention strategy in atrial fibrillation (AF) across all chronic kidney disease (CKD) stages.

**Methods:** We used the Cockcroft-Gault (CG), Modified Diet in Renal Disease (MDRD), and Chronic Kidney Disease Epidemiology Collaboration (CKD-EPI) equations to classify 39,217 patients into stage 1 to 5 CKD during July 1^st^, 2001, and September 30^st^, 2018. The endpoint is a composite outcome including ischemic stroke or major bleeding or mortality.

**Results:** More patients belonged to stage 1 and 2 CKD using the MDRD and CKD-EPI equations. In subgroups of patients with eGFR-MDRD or eGFR-CKD-EPI ≥ 60 mL/min, a 17-18% increase of event was observed in patients with eGFR-CG < 60 mL/min compared to those ≥ 60 mL/min. Compared to no oral anticoagulant (OAC), OAC use was associated with a significantly lower risk of event across stage 1 to 4 CKD but not in stage 5 CKD. Both warfarin and NOACs exhibited better outcome compared to no OAC across stage 1 to 4 CKD while NOACs was associated with more risk reduction compared to warfarin. Among patients on OACs, there was a trend toward better outcome with NOAC than warfarin across stage 2-4 CKD but not in stage 1 and 5 CKD.

**Conclusions:** OAC should be used in stage 1 to 4 CKD with NOAC exhibiting the trend of better outcome through stage 2 to 4 CKD than warfarin. For stage 5 CKD, optimal strategy remains undetermined.

**Clinical Perspective:**

- **What Is New?** The stages of renal function of AF patients varied significantly with different renal equations, and tthe CG equation remained effective in differentiating clnical outcomes for patients with eGFR-MDRD ≥ 60 mL/min or eGFR-CKD-EPI ≥ 60 mL/min
- **What Are the Clinical Implications?** OAC should be used in stage 1 to 4 CKD with NOAC exhibiting the trend of better outcome through stage 2 to 4 CKD than warfarin.

## Introduction

Atrial fibrillation (AF) is the most common cardiac arrhythmia worldwide and increases the risk of ischemic stroke, cardiovascular events, and all-cause mortality.^1^ Hypercoagulability and the irregular hemodynamic status of AF place patients vulnerable to renal function impairment,^2^ so chronic kidney disease (CKD) is common in patients with AF.^3, 4^ On the other hand, renal function impairment increases the risk of ischemic stroke and major bleeding in AF.^5–7^ For example, stroke rates increased from 1.4% per year in stage 1 CKD to 3.72% in stage 4-5 CKD, and so did the rates of mortality and bleeding.^8^

Oral anticoagulants (OACs) are indicated in AF patients at high risk of ischemic stroke after weighing the risk and benefit of OACs.^1, 9^ Non-vitamin K antagonist (NOAC) is the mainstream OACs in stroke prevention in current guidelines because of better safety and comparable or even better efficacy compared to warfarin.^1, 9–13^ However, because different types of NOACs undergo certain degree of renal clearance, patients with severe CKD were mostly excluded from landmark randomized controlled trials (RCTs).^14–17^ Besides, there are conflicting results for the use of NOACs and warfarin in patients with end-stage CKD and calciphylaxis is a major concern with the use of warfarin in end-stage CKD.^18, 19^ Therefore, the use of OAC in AF patients with CKD remains challenging. Although there were real-world cohort studies specifically analyzed OAC use in subgroups of patients with advanced CKD,^20–26^ a simultaneous comparison of OAC strategies across all CKD stages is lacking. Therefore, the aim of the present study is to use a large-scale real-world cohort to simultaneously analyze the effect of OAC use and OAC types in AF patients across all stages of CKD. Furthermore, how different renal function equations affect the classification of CKD stage and its role in differentiating prognosis will also be studied.

## Methods

### Database

This is a retrospective study using the data from the Chang Gung Research Database provided by Chang Gung Memorial Hospital (CGMH) Medical system. The CGMH Medical system included 3 major teaching hospitals and 4 tertiary care medical centers with a total of 10,050 beds and about 280,000 admissions per year.^27^ There were around 500,000 emergent department visits and 8,500,000 outpatient department visits to CGMH Medical system in 2015, accounting for approximately 1/10 of the annual Taiwanese medical service.^27^ In this database, the personal information and identification number of each patient are encrypted and a consistent encrypting procedure was used to de-identify detailed medical information of each patient.^28^ Therefore, informed consent was waived for this study. This study was approved by the Institutional Review Board of the Chang Gung Medical Foundation (201802075B0).

### Study cohort and study design

Patients ≥20 years with newly diagnosed AF during July 1^st^, 2001, and September 30^st^, 2018, were identified from the CGMH medical database (n = 70,408) and we excluded patients without body weight (BW) and serum creatinine (sCr) data within 6 months before AF being diagnosed. A total of 39,217 patients were ultimately included for the present study.

### Renal function determination

The estimated glomerular filtration rate (eGFR) was calculated for renal function evaluation using the Cockcroft-Gault (CG), Modified Diet in Renal Disease (MDRD), and Chronic Kidney Disease Epidemiology Collaboration (CKD-EPI) equations as the following:^29–31^

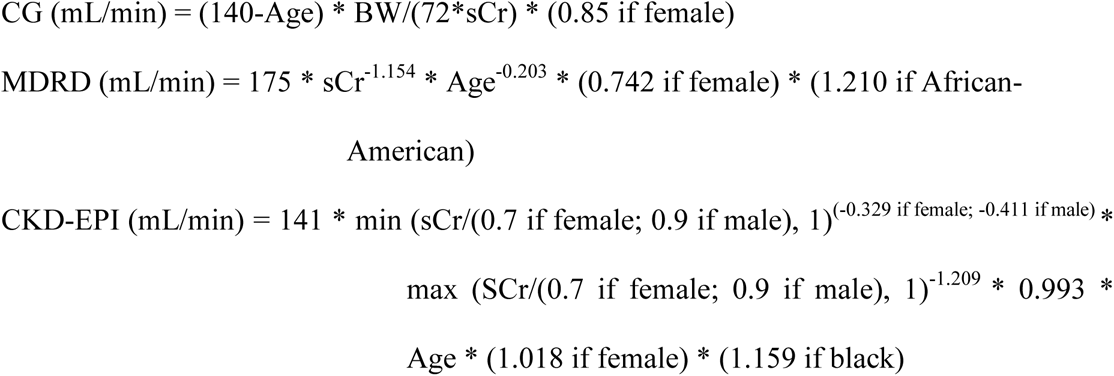

Patients were classified as stage 1 CKD (eGFR > 90 mL/min), stage 2 CKD (eGFR of 60 to 89 mL /min), stage 3 CKD (eGFR of 30 to 59 mL /min), stage 4 CKD (eGFR of 15 to 29 mL /min), and stage 5 CKD (eGFR < 15 mL/min).

### Clinical endpoints

The clinical endpoint is a composite outcome including the occurrence of ischemic stroke or major bleeding or all-cause mortality. A stratified analysis comparing OAC strategy and OAC types across all CKD stages was performed. All study endpoints were defined based on the first discharge diagnosis to avoid misclassification. Major bleeding was defined as hospitalization of intracranial hemorrhage, gastrointestinal bleeding, and other sites of critical bleeding. The follow-up period was defined as the duration from the date when AF was diagnosed until the occurrence of clinical outcomes, or until the end date of the study period (September 30, 2018), whichever came first.

### Statistical analysis

Data are presented as the mean value ± standard deviation for continuous variables and proportions for categorical variables. Differences between continuous values were assessed using the unpaired two-tailed *t*-test or one-way analysis of variance (ANOVA). Differences between nominal variables were compared by the chi-squared test. The incidences of clinical endpoint were calculated from dividing the number of events by person-time at risk. The risk of clinical endpoint was assessed using the Cox regression analysis. The proportional hazards assumption was tested using Schoenfeld residual test which showed no non-proportionality. The Kaplan-Meier method was used to plot the cumulative incidence curves of clinical endpoints for different renal function groups, with statistical significance examined by the log-rank test. All statistical significances were set at a *p* < 0.05.

## Results

### Baseline characteristics of all AF patients

**Table 1** shows the baseline characteristics of the study population. The mean age of the 39,217 patients with AF was 71.09 ± 12.76 years and 57.1% of them were males. The mean CHA_2_DS_2_-VASc and HAS-BLED scores were 2.58 ± 1.65 and 1.94 ± 1.33, respectively. Hypertension was the most common comorbidity with 47.8% of patients having hypertension. The mean and median sCr levels were 1.62 ± 1.91 mg/dL and 1.03 mg/dL (interquartile range 0.80-1.45 mg/dL), respectively. The mean eGFRs were 56.75 mL/min calculated by CG equation, 69.75 mL/min by MDRD equation and 62.12 mL/min by CKD-EPI euqation. Among all 39,217 patients, 12,791 patients were treated with OAC, including 6,381 patients on warfarin and 6,410 patients on NOACs.

**Table 1.** Baseline characteristic of all AF patients.

|  | **All (n=39,217)** |
| --- | --- |
| **Age (years), mean (SD)** | 71.09 (12.76) |
| **Age≥75 years, n (%)** | 10545 (26.9) |
| **Age 65-74 years, n (%)** | 17634 (45.0) |
| **Sex (male), n (%)** | 22374 (57.1) |
| **Body weight, kg** | 63.15 (13.76) |
| **CHA<sub>2</sub>DS<sub>2</sub>-VASc score, mean (SD)** | 2.58 (1.65) |
| **HAS-BLED score, mean (SD)** | 1.94 (1.33) |
| **Comorbidities, n (%)** |  |
| **Hypertension** | 18755 (47.8) |
| **Diabetes mellitus** | 9918 (25.3) |
| **Heart failure** | 1340 (3.4) |
| **Prior stroke/TIA** | 2592 (6.6) |
| **Vascular diseases** | 2953 (7.5) |
| **COPD** | 9082 (23.2) |
| **Hyperlipidemia** | 9396 (24.0) |
| **Serum Creatinine level (mg/dL), mean (SD)** | 1.62 (1.92) |
| **Serum Creatinine level (mg/dL), median (IQR)** | 1.03 (0.80 – 1.45) |
| **eGFR by CG equation (mL/min)** | 56.75 (34.68) |
| **eGFR by MDRD equation (mL/min)** | 69.75 (39.94) |
| **eGFR by CKD-EPI equation (mL/min)** | 62.12 (29.36) |
| **Stroke prevention strategy, n (%)** |  |
| **Non-OACs** | 26426 (67.4) |
| **Warfarin** | 6381 (16.3) |
| **NOACs** | 6410 (16.3) |

AF = atrial fibrillation; COPD = chronic obstructive pulmonary disease; eGFR = estimated glomerular filtration rate; IQR = interquartile range; NOACs = non-vitamin K antagonist oral anticoagulant; SD = standard deviation; TIA = transient ischemic attack

### Varied renal function classification based on different equations and its relationship with clinical endpoint

Patient classifications based on renal function status were dissimilar based on different equations. The CG equation classified patients into 14.7% of stage 1 CKD, 25.9% of stage 2 CKD, 37.2% of stage 3 CKD, 12.7% of stage 4 CKD, and 9.4% of stage 5 CKD, respectively. With MDRD equation, there were 24.5% of stage 1 CKD. 35.5% of stage 2 CKD, 25.3% of stage 3 CKD, 6.7% of stage 4 CKD, and 8.1% of stage 5 CKD. Based on the CKD-EPI equation, the distribution of renal function stages was as follows: 18.0% of stage 1 CKD, 37.8% of stage 2 CKD, 27.7% of stage 3 CKD, 7.6% of stage 4 CKD, and 8.9% of stage 5 CKD. In brief, part of patients with eGFR < 60 mL/min based on the CG equation might have been classified as stage 1 or 2 CKD using MDRD or CKD-EPI equations. (**Figure 1**) In subgroup of patients with eGFRs-MDRD or eGFRs-CKD-EPI ≥ 60 mL/min, patients with eGFR-CG < 60 mL/min was significantly associated with 17% and 18% increase of clinical endpoint compared to those with eGFR-CG ≥ 60 mL/min. (**Figure 2**)

**Figure 1.**
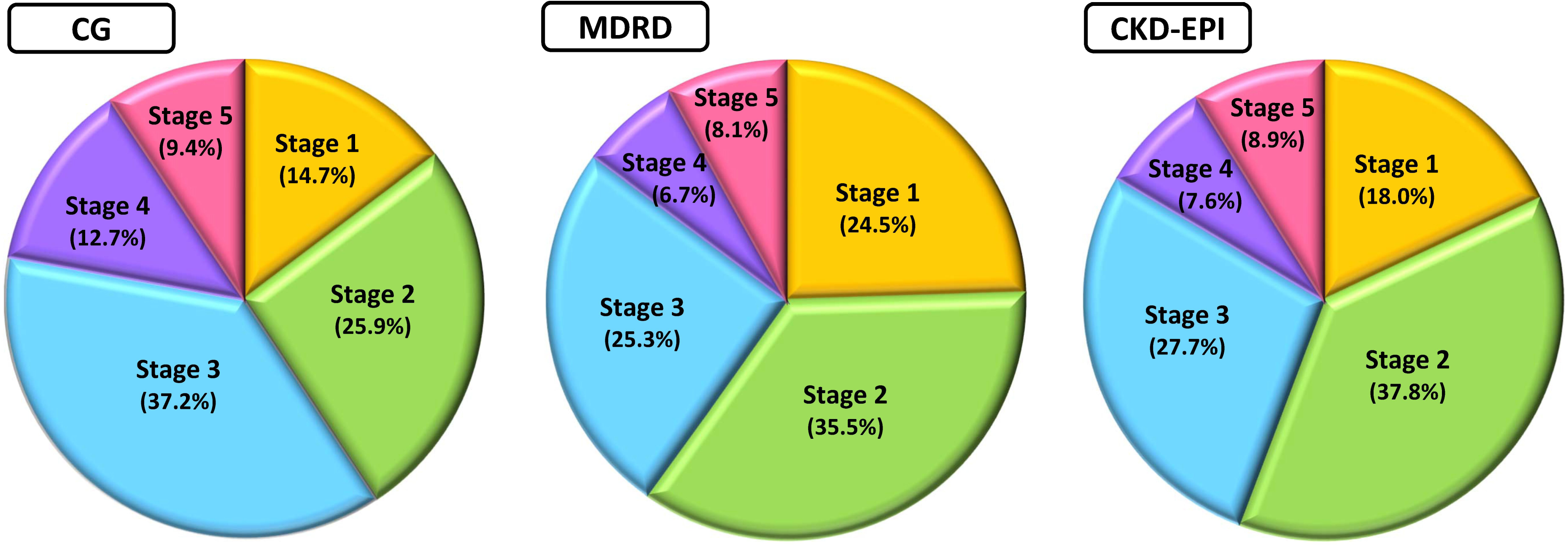
The distribution of renal function stages using different equations. These pie charts illustrated the distribution of renal function stages using the CG, MDRD, and CKD-EPI equations. More patients were classified as stage 1 and 2 CKD using the MDRD and CKD-EPI equations than the CG equation, which highlighted the discrepancy of renal function classification between different equations.

**Figure 2.**
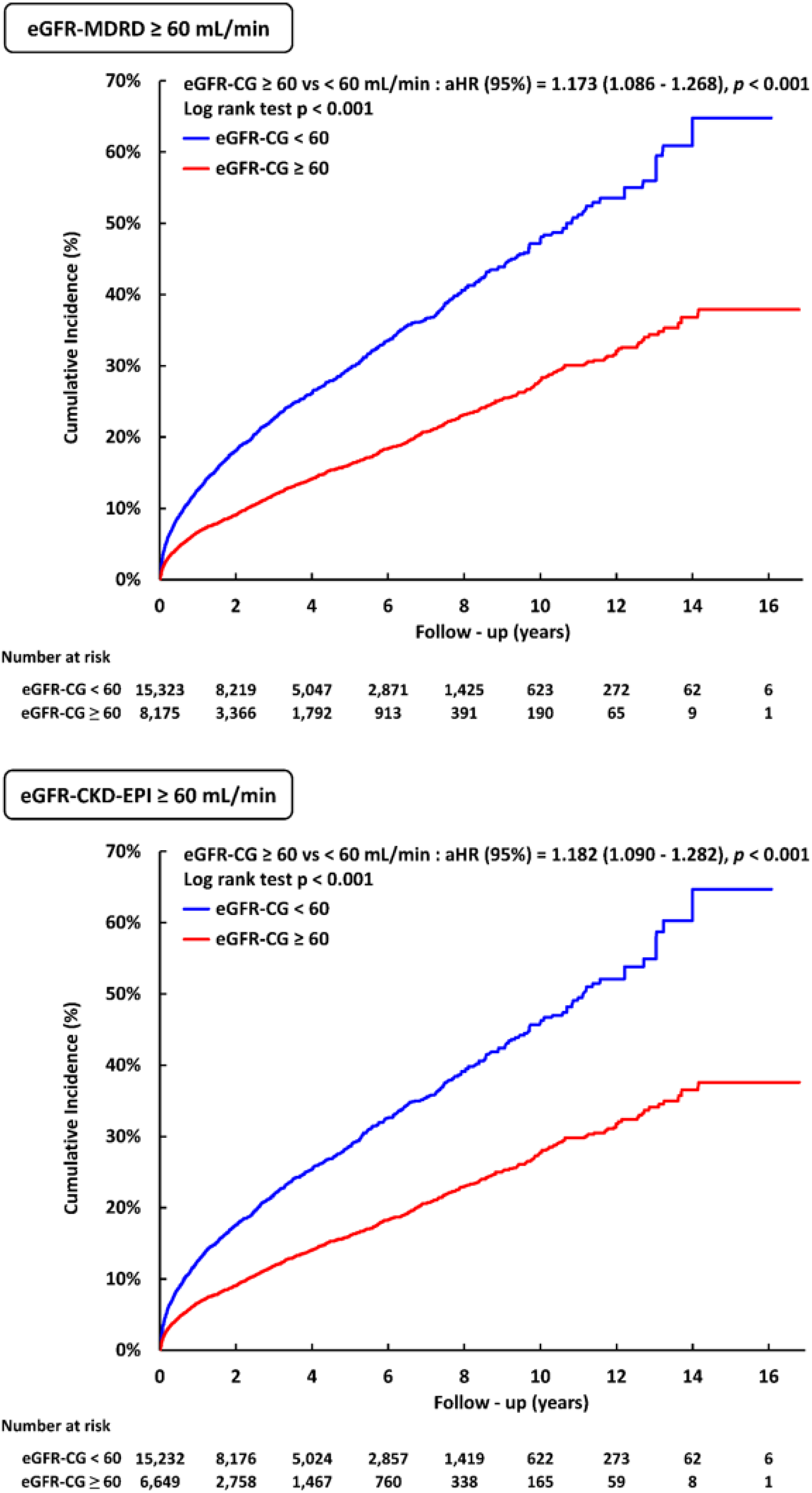
Cumulative incidence of clinical events in patients with eGFR-MDRD and eGFR-CKD-EPI ≥ 60 mL/min. The Kaplan-Meier method disclosed that even in patients with eGFR above 60 mL/min using the MDRD or CKD-EPI method, the CG equation with a cut-off value of 60 mL/min still differentiated patients into different prognosis. Those with eGFR-CG < 60 mL/min was associated with worse outcome compared to patients with eGFR-CG ≥ 60 mL/min. aHR = adjusted hazard ratio; eGFR = estimated glomerular filtration rate

### The impact of OAC use on clinical endpoint in each CKD stage

Among all study population, the use of OAC was associated with 29.7% decrease of clinical endpoint compared to those without OACs, and more risk reduction was observed on NOACs than warfarin (NOACs: adjusted hazard rate [aHR]: 0.497, 95% confidence interval [CI]: 0.456-0.543; warfarin: aHR 0.863, 95% CI 0.810-0.919; interaction P < 0.001). This was also evident through stage 1 to 4 CKD but not in stage 5 CKD. (**Figure 3**) There was no significant difference of risk between OAC and no OAC groups in patients with stage 5 CKD.

**Figure 3.**
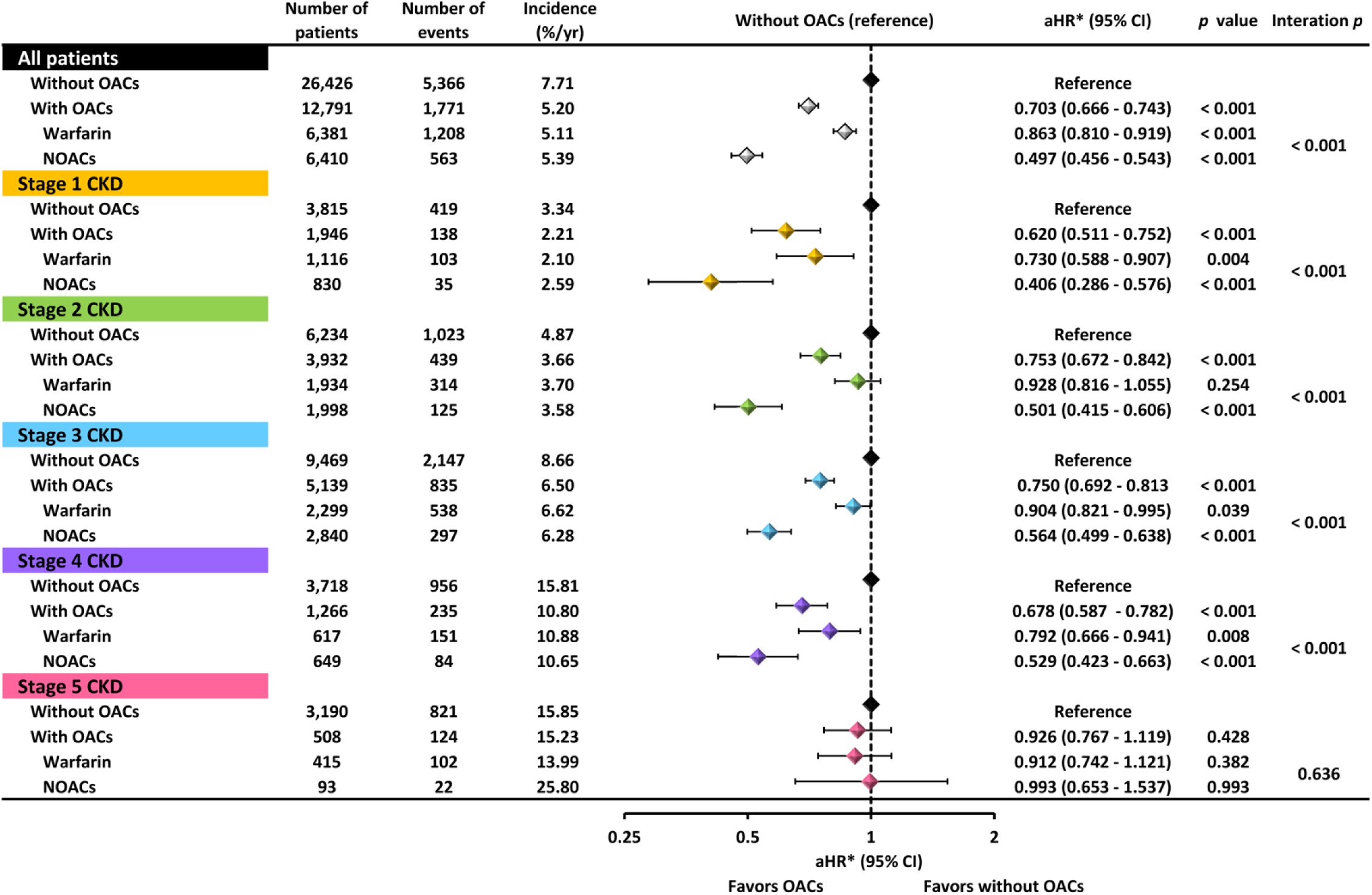
Risk of clinical events across all stages of CKD. Compared to no OAC, OAC use was associated with better outcomes in stage 1-4 CKD whereas NOACs exhibited more risk reduction than warfarin compared to no OAC. In stage 5 CKD, there was no differences between OAC use or not. aHR = adjusted hazard ratio; CI = confidence interval; CKD = chronic kidney disease; NOACs = non-vitamin K antagonist oral anticoagulants; OACs = oral anticoagulants

Subgroup analysis in patients on OAC in each CKD stage was performed. (**Figure 4**) In patients with stage 1 and 5 CKD, there was no significant difference of the risk of clinical endpoint between NOAC and warfarin (stage 1 CKD: aHR 0.937, 95% CI 0.607-1.447, P = 0.768; stage 5 CKD: aHR 1.168, 95% CI 0.711-1.919, P = 0.539). In patients with stage 2 and stage 3 CKD, multivariate Cox regression analysis demonstrated significantly lower risk with NOAC compared to warfarin. (stage 2 CKD: aHR 0.630; 95% CI 0.504-0.792; stage 3 CKD: aHR 0.712, 95% CI 0.610-0.832; both P < 0.001). In stage 4 CKD, NOAC was associated with a trend toward lower risk, although a marginal P value of 0.052 was observed. (**Figure 4**)

**Figure 4.**
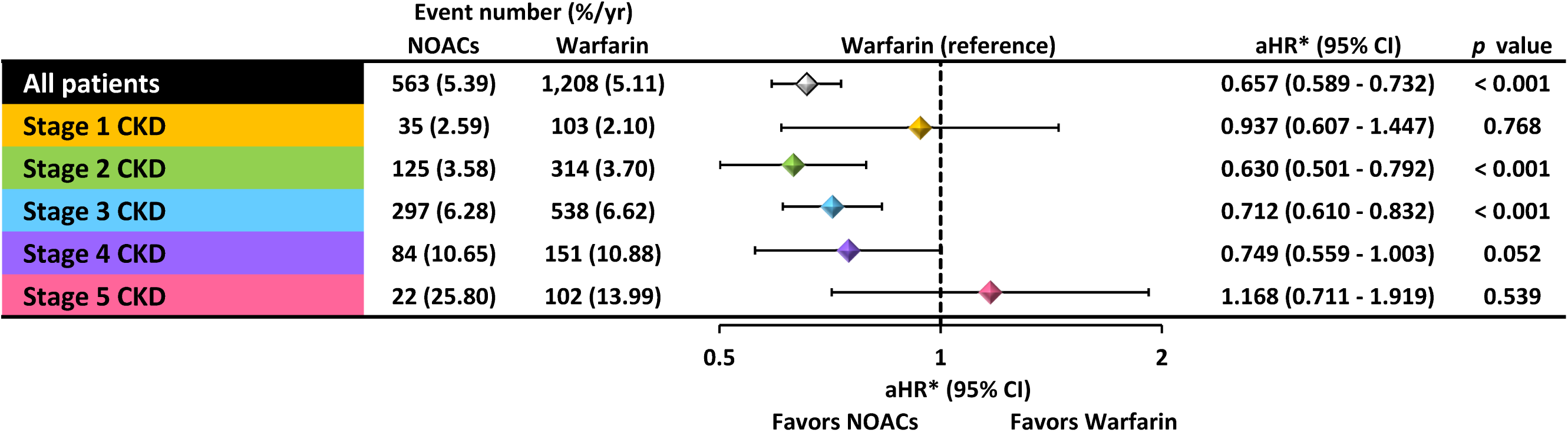
Subgroup analysis comparing warfarin with NOACs in patients receiving oral anticoagulants. Compared to warfarin, NOAC was associated with significantly lower risk of clinical events in stage 2-3 CKD while there was a marginal benefit toward NOAC use in stage 4 CKD. In patients with stage 1 and 5 CKD, however, there was no significant differences between warfarin and NOACs. aHR = adjusted hazard ratio; CI = confidence interval; CKD = chronic kidney disease; NOACs = non-vitamin K antagonist oral anticoagulants

## Discussion

### Main findings

In the present study, a large Asian AF cohort was used to investigate the risk of clinical outcomes in relation to the evaluation of renal function status and OAC strategy. The main findings are as follows: (i) patient grouping varied dependidng on the renal function equations used, and the CG equation was effective in differentiating clnical outcomes for patients with eGFR-MDRD ≥ 60 mL/min or eGFR-CKD-EPI ≥ 60 mL/min; (ii) compared to no OAC, OAC use was associated with significantly lower risk of clinical events through stage 1 to 4 CKD but not in stage 5 CKD; and (iii) among patients with stage 2 to 4 CKD, NOAC was associated with better outcomes than warfarin, while there was no significant difference between NOAC and warfarin in patients with stage 1 and 5 CKD.

### The role of renal function equations in differentiating clinical outcomes

The CG equation, which considers patient’s age, gender, and body weight, was mostly used to estimate patient’s renal function in all landmark NOAC trials and international guidelines,^32–34^ whereas the MDRD and CKD-EPI equations are frequently used in real-world practice. There were studies demonstrating inconsistence of renal function estimation using different equations in AF patients.^35–38^ Misclassification of renal function status in AF patients using the MDRD and CKD-EPI equations was mostly observed,^37, 38^ and may have harmful effect on prognosis because of potential off-label dosing of NOACs.^37^ Therefore, the CG equation is recommended for correct dosing of NOACs.^38, 39^ Our present study also observed disagreement of renal function classification according to different equations. In subgroup of patients with preserved renal function as presented with eGFR-MDRD ≥ 60 mL/min or eGFR-CKD-EPI ≥ 60 mL/min, a cut-off value of eGFR-CG of 60 mL/min still differentiated patients into different outcomes. Therefore, our study again reinforces that CG equation is preferred over MDRD and CKD-EPI equations for renal function estimation and determining NOAC doses in AF patients.

### The effect of OAC on clinical outcomes in each renal function stage

To the best of our knowledge, there is no study simultaneously analyzing the stroke prevention strategy in each CKD stage in AF patients. Only scattered cohort studies or subgroup analysis aiming at certain OAC or particular CKD stages were available. Sub-analysis in patients with stage 3 to 4 CKD from large-scale RCTs showed that NOACs were non-inferior to warfarin in stroke prevention with comparable or less risk of bleeding.^20–24^ A meta-analysis including 15 studies in patients with mild, moderate, and severe CKD observed no difference between NOAC and warfarin in the risk of storke or systemic embolism in any subgroups of CKD. NOAC further reduced the risk of mortality in moderate-severe or severe CKD and major bleeding in modeate and moderate-severe CKD.^40^ A retrospective study including 21,733 AF patients with CKD reported NOAC being associated with lower mortality, less bleeding, and a non-significant trend toward lower embolic stroke compared to warfarin in all renal function groups.^41^ In the present study, we displayed the analysis in an organized manner covering all CKD stages, and clearly demonstrated the benefit of OAC through stage 1-4 CKD but not stage 5 CKD. NOACs especially showed less risk through stage 2-4 CKD but not in stage 1 and 5 CKD. We suppose that the prognosis of stage 1 CKD might be too good to tell the differences of prognosis between NOAC and warfarin. As to stage 5 CKD which was universally excluded from large-scale RCTs,^42–45^ we found no difference of risk between OAC use or not and between NOAC and warfarin in stage 5 CKD, which further highlights the dilemma of stroke prevention in AF patients with end-stage CKD.

In brief, although there is a long way to go before more robust data are available, our findings are in line with a review from a nephrological perspective in which they proposed NOACs to be the first line therapy for AF patients with mild and moderate renal dysfunction and may be adopted for those with severe CKD not on dialysis.^46^ For stage 5 CKD, limited studies suggested apixaban use, but overall, the benefit of OAC or NOACs in AF patients with stage 5 CKD remain an uncharted territory.

### Limitations

There are several limitations of the present study. First, this was a retrospective study of the electronic medical record database, so the decision of OAC was not per pre-specified algorithm and depended on the physicians who were in charge of those patients. Therefore, conditions precluding or preferring/inclining a certain strategy may be present. Second, although we used multivariable Cox regression analysis to adjust baseline differences, some unmeasured confounders may still exist. Third, the estimation of renal fucntion stages were based on the data and BW at enrollment. We can not exlcude the possibility of fluctuation of renal function during the follow-up period and its impact on prognosis. Fourth, the time in therapeutic range of warfarin use was not available in the present study, which might influence the effect of warfarin. Last, individual NOAC use and its dosing in different renal function status were not included in the analysis. So it is unknown whether one NOAC is superior than the other in distinct renal function stages.

## Conclusion

The stages of renal function of AF patients varied significantly with different renal equations. OAC use was associated with better prognosis than no OACs through stage 1 to 4 CKD, whereas NOACs was better or at least non-inferior to warfarin through stage 2 to 4 CKD. For stage 5 CKD, optimal strategy of stroke prevention, including the use of OAC or not and OAC types, remains unknown, and more studies are warranted.

## Data Availability

The data underlying this article are available in the article.

## Acknowledgments

The authors thank the statistical assistance and wish to acknowledge the support of the Maintenance Project of the Center for Big Data Analytics and Statistics (Grant CLRPG3D0045) at Chang Gung Memorial Hospital for study design and monitor, data analysis and interpretation.

